# Assay-dependent Effects of EDTA Contamination on Plasma Magnesium and Iron

**DOI:** 10.1101/2025.03.06.25323264

**Authors:** Davor Brinc, Osmond Rodrigues, Fari Rokhforooz, Felix Leung, Dana Nyholt, Rajeevan Selvaratnam

**Author notes:** **Corresponding author:** Rajeevan Selvaratnam, Ph.D., Division of Clinical Biochemistry, Laboratory Medicine Program, University Health Network, Toronto, Canada, Department of Laboratory Medicine & Pathobiology, University of Toronto, Toronto, ON, Canada.

## Abstract

**Background:** Inadvertent submission of ethylenediaminetetraacetic acid (EDTA) based plasma in place of heparinized plasma or contamination of serum or heparinized plasma with EDTA can alter select laboratory measurements. While factitious hypocalcaemia and abnormally low alkaline phosphatase (ALP) are recognized indicators of EDTA contamination, the impact of EDTA on Mg^2+^ and Fe^2+^ remains controversial.

**Method:** Herein, we derived the EDTA concentration required to cause a 50% decline (EC50_[EDTA]_) for Ca^2+^, ALP, Mg^2+^, and Fe^2+^ across two methodologies (Roche Cobas c303 and Abbott Alinity *c*) by varying EDTA concentration across plasma and serum pools. The concentration of EDTA required to exceed the allowable performance limit (APL_[EDTA]_) was also evaluated across methods.

**Results:** Mg^2+^ measured by an isocitrate dehydrogenase enzymatic method was resilient against EDTA, while Mg^2+^ measured by xylidyl blue had an EC50_[EDTA]_ and APL_[EDTA]_ of 0.78 – 1.18 mmol/L and 0.16 – 0.34 mmol/L, respectively. Fe^2+^ measured with a ferene method was resilient to EDTA, whereas ferrrozine method indicated EC50_[EDTA]_ and APL_[EDTA]_ of 5.6 – 8.86 mmol/L and 4.68 – 5.46 mmol/L, respectively. Ca^2+^ and ALP exhibited larger EC50_[EDTA]_ on the Abbott Alinity *c*.

**Conclusion:** Hypomagnesemia and hypoferremia are not definitive markers of potential EDTA contamination, as Mg^2+^ and Fe^2+^ measurements show method-dependent susceptibilities.

## Introduction

Ethylenediaminetetraacetic acid (EDTA) contamination is a pre-analytical error that has been characterized as having a noticeable impact on several clinical chemistry measurands. For example, it has been well established that such contamination leads to a false increase in K^+^ (as EDTA is introduced as a potassium salt), decrease in Ca^2+^ and Mg^2+^ from chelation by EDTA, and a consequent decrease in alkaline phosphatase (ALP). Davidson et al. illustrated this classical phenomenon of concomitant depression of Ca^2+^ and Mg^2+^ in the presence of K_3_EDTA contamination, highlighting the chelation effects on clinical measurements(1). While the effect of EDTA on measured Ca^2+^ and ALP activity has been consistent across studies(1–4), the effect on measured Mg^2+^ is conflicting. Some studies have reported significant declines in Mg^2+^ from EDTA contamination (1,4–6) while others have shown no significant effect on measured Mg^2+^ (2,3). In particular, the latter studies utilized the Abbott platforms(2,3) with methodologies for Mg^2+^ being an enzymatic method, whereas the former studies illustrating notable declines in Mg^2+^ with EDTA contamination (1,4–6) all relied on the Roche platform, which utilizes a dye binding method (xilidyl blue).

Similarly, several studies have implicated a notable decline in Fe^2+^ measurement in the presence of EDTA(1,5,7,8) with the method being the reaction of liberated Fe^2+^ with ferrozine on the Roche platforms. Only one recent study to our knowledge implies limited reportability of Fe^2+^, with the method being on the Abbott Architect(2), which uses ferene as the iron chromogen(9). Structurally, both ferene and ferrozine behave as bidentate chelating agents, with the only structural difference between ferrozine and ferene being a substitution of an oxygen atom in each of the furyl rings in ferene (10).

Previous studies evaluating the EDTA effects have looked at only a single method for assessing impact and have implicated hypomagnesemia and hypoferremia as surrogate markers of EDTA contamination. However, EDTA may not always elicit factitious hypomagnesemia and hypoferremia. Here we evaluated the EDTA effects on two commonly used methods for measuring not only Mg^2+^ and Fe^2+^, but also Ca^2+^ and ALP on the Abbott Alinity *c* and the Roche cobas c303. We illustrate the method dependent differences by modelling the decline for each surrogate marker of EDTA contamination and determining the effective range of EDTA concentration required to cause a 50% decline from baseline (EC50_[EDTA]_) as well as the EDTA concentration required to exceed the allowable performance limit (APL_[EDTA]_).

## Materials and Methods

A total of four sample pools, designated as Sample 1-4, were generated from residual plasma or sera. Samples 1-3 were obtained from collections in plasma separator tubes containing lithium heparin (Beckton, Dickinson and Company, NJ, USA) and Sample 4 was obtained from a pool of collections obtained from serum separator tubes (Beckton, Dickinson and Company, NJ, USA). All sample pools were free of hemolysis and lipemia. All sample pools except Sample 3 were free of icterus, with Sample 3 having a 3+ or an average of 293 μmol/L of icterus as determined on the Abbott Alinity *c* instrument. This level of icterus was below the interference threshold for all assays except ALP2 on Abbott Alinity *c*, with manufacturer claim of 178 μmol/L and 342 μmol/L of conjugated and unconjugated bilirubin, respectively, as the icteric threshold. A stock concentration of Na_2_-EDTA (Caledon Laboratories Ltd, Georgetown, ON Canada) was generated and spiked into seven aliquots of Samples 1-4, with a final EDTA concentration of 0.10, 0.25, 0.50, 1.01, 2.01, 4.02, and 8.05 mM. An aliquot of Samples 1-4 without EDTA spiking served as baseline. Matrix alteration of all samples including baseline, as a result of spiking was 8% (v/v).

All sample aliquots were assayed on the Abbott Alinity *c* for ALP, Mg^2+^, and Fe^2+^. These aliquots were subsequently assayed on the Roche cobas c303 method for the aforementioned tests. The ALP method is version 2 (ALP2) of the Abbott method using a factor-based calibration (1931) (ref: 04T83). Both the Abbott and Roche ALP2 method (ref: 08056757190) have claimed traceability to the IFCC reference method. Mg^2+^ measurement on the Abbott Alinity *c* was based on the enzymatic (isocitrate dehydrogenase) method (ref: 08P19), whereas the Roche cobas c303 was based on the xylidyl blue dye binding method (ref: 06407358190, 06407358214). Ca^2+^ was determined by a dye binding method, Arsenazo-III on the Abbott Alinity *c* (ref: 07P57) and by a chelation method *via* 5□nitro□5’□methyl□BAPTA (NM-BAPTA) and EDTA on Roche cobas c303 (ref: 08057427190, 08057427214). Fe^2+^ was determined by acidification to liberate transferrin bound Fe^3+^, reduction with ascorbate to Fe^2+^, and then reaction with ferene (Iron2 reagent on Abbott Alinity *c*, ref: 04T98) or ferrozine (Iron2 reagent on the Roche cobas c303, ref: 08057931190). For method comparison, 20 residual patient samples were evaluated on the Abbott Alinity *c*, and subsequently analyzed on the Roche cobas c303 on the same day.

All statistical, regression, and graphical analyses were performed in Python [version 3.12.4]. The SciPy [version 1.15.0] module was used for curve fitting a Hill-Langmuir equation to model a dose-response function and for deriving the effective concentration of EDTA required to decrease the baseline concentration of analyte by 50% (*i*.*e*., EC50_[EDTA]_). The EDTA concentration required to breach the allowable performance limits (*i*.*e*, APL_[EDTA]_) from a given baseline concentration was back calculated using the derived EC50_[EDTA]_. Allowable performance limits were obtained from Institute of Quality Management in Health Care, a subsidiary of Accreditation Canada Diagnostics and were as follows: ± 0.10 mmol/L at < 1.25 mmol/L of Mg^2+^, ± 0.12 mmol/L at <2.50 mmol/L Ca^2+^, ± 3 μmol/L at < 20 μmol/L Fe^2+^, ± 15 U/L at < 100 U/L of ALP, and ± 15 % at ≥ 100 U/L of ALP. All plots were generated using the Plotly module [version 5.23] in Python.

## Results

Across four sample pools, Mg^2+^ measured by the xilidyl blue dye method led to a rapid decrease with increasing concentration of EDTA (**Figure 1**, top panel). Specifically, the EC50_[EDTA]_ for Mg^2+^ ranged from 0.78 to 1.18 mmol/L of EDTA whereas APL_[EDTA]_ ranged from 0.16 – 0.34 mmol/L (**Table 1**). At the highest concentration of EDTA, there was little to no measurable Mg^2+^ with the dye method (**Figure 1**, top panel). The enzymatic measurement of Mg^2+^ by the Abbott Alinity *c* had very modest decline, most evident at the highest concentration of EDTA, but within in APL (**Supplementary Figure S1**). In the absence of EDTA, the Mg^2+^ measurements were highly correlated (Pearson’s R = 0.999) with a median bias of 0.048 mmol/L relative to the enzymatic method (**Supplementary Figures S2**).

**Figure 1.**
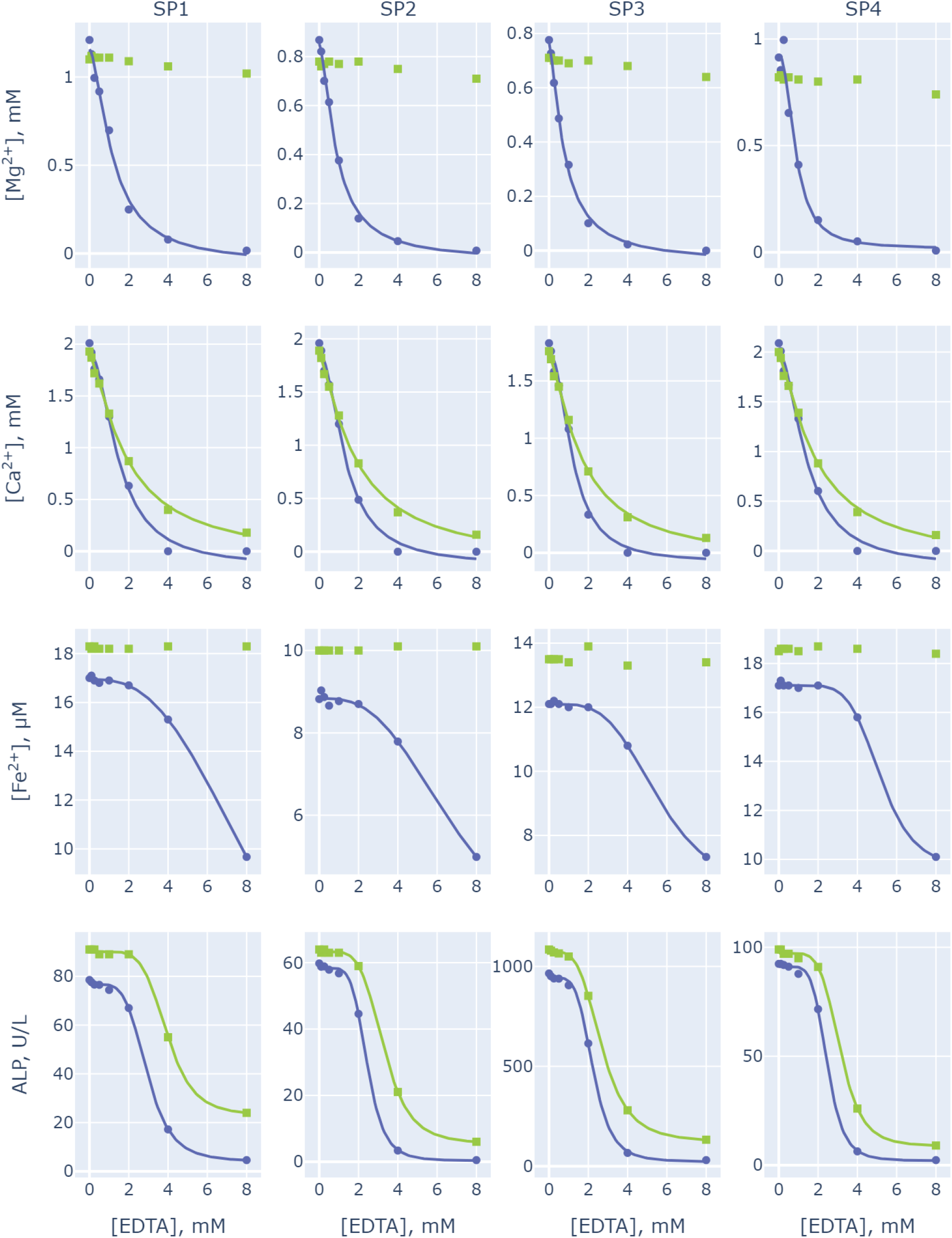
The effect of EDTA concentration on Mg^2+^ (row 1), Ca^2+^ (row 2), Fe^2+^ (row 3) and ALP activity (row 4) as determined by two methods, Abbott Alinity *c* (green square) and Roche cobas c303 (violet circles). Each column represents a different Sample Pool (SP) 1 – 4 as described in the text.

**Table 1.** EC50_**[EDTA]**_ **and APL**_**[EDTA]**_ **for Mg**^**2+**^, **Ca**^**2+**^, **Fe**^**2+**^, **and ALP**

| Analyte | Method | SP 1 |  |  | SP 2 |  |  | SP 3 |  |  | SP 4 |  |  |
| --- | --- | --- | --- | --- | --- | --- | --- | --- | --- | --- | --- | --- | --- |
|  |  | [Baseline] | EC50<sub>[EDTA]</sub><br>(mmol/L) | APL<sub>[EDTA]</sub><br>(mmol/L) | [Baseline] | EC50<sub>[EDTA]</sub><br>(mmol/L) | APL<sub>[EDTA]</sub><br>(mmol/L) | [Baseline] | EC50<sub>[EDTA]</sub><br>(mmol/L) | APL<sub>[EDTA]</sub><br>(mmol/L) | [Baseline] | EC50<sub>[EDTA]</sub><br>(mmol/L) | APL<sub>[EDTA]</sub><br>(mmol/L) |
| Mg<sup>2+</sup><br>(mmol/L) | Xylidyl Blue,<br>Roche | 1.21 | 1.18 | 0.16 | 0.87 | 0.87 | 0.19 | 0.78 | 0.78 | 0.16 | 0.91 | 0.86 | 0.34 |
|  | Isocitrate<br>dehydrogenase,<br>Abbott | 1.10 | - | - | 0.78 | - | - | 0.71 | - | - | 0.82 | - | - |
| Ca<sup>2+</sup>,<br>(mmol/L) | Chelation NM-<br>BAPTA, Roche | 2.01 | 1.49 | 0.16 | 1.96 | 1.31 | 0.18 | 1.83 | 1.20 | 0.19 | 2.09 | 1.42 | 0.15 |
|  | Arsenazo III,<br>Abbott | 1.93 | 1.86 | 0.19 | 1.89 | 1.86 | 0.17 | 1.76 | 1.67 | 0.18 | 2.00 | 1.89 | 0.17 |
| Fe<sup>2+</sup><br>(μmol/L) | Ferrozine,<br>Roche | 17.0 | 8.86 | 4.75 | 8.8 | 7.2 | 5.46 | 12.1 | 5.6 | 4.68 | 17.1 | 8.68 | 5.08 |
|  | Ferene, Abbott | 18.3 | - | - | 10.0 | - | - | 13.5 | - | - | 18.5 | - | - |
| ALP,<br>(U/L) | Enzymatic,<br>Roche | 79 | 2.93 | 2.06 | 60 | 2.44 | 1.90 | 965 | 2.25 | 1.49 | 92 | 2.46 | 1.82 |
|  | Enzymatic,<br>Abbott | 91 | 3.94 | 3.02 | 64 | 3.28 | 2.49 | 1085 | 2.69 | 1.70 | 99 | 3.12 | 2.24 |

Measurements of Ca^2+^ were higher using NM-BAPTA (Roche) versus Arsenazo-III (Abbott) (**Supplementary Figure S3**; median bias 0.10 mmol/L) but were well correlated (Pearson’s R = 0.992). Both methods were susceptible to EDTA interference, with steeper declines for the NM-BAPTA method (**Figure 1**, second row panels). This is also evident by lower EC50_[EDTA]_ values (1.20 – 1.49 mmol/L) compared to the Abbott’s Arsenazo-III dye method (1.67 – 1.89 mmol/L). The APL_[EDTA]_ for Ca^2+^ measurement with NM-BAPTA and Arsenazo-III were similar, ranging from 0.15 – 0.19 mmol/L and 0.17 – 0.19 mmol/L, respectively, across pools (**Table 1**).

ALP activity measures were proportionally lower with the Abbott method and in the absence of EDTA, but with high degree of correlation (Pearson’s R = 1.0) between both methods (**Supplementary Figure S4**). The effect of EDTA on both the Roche and Abbott assays for ALP activity were similar, leading to an overall saturable loss of activity by both methods (**Figure 1**, bottom panel). The EC50_[EDTA]_ values for ALP activity were lower with the Roche method (2.25 – 2.93 mmol/L) in comparison to Abbott (2.69 – 3.94 mmol/L).

The ferrozine method for Fe^2+^ measurement by the Roche method was observed to have a more significant decrease in an EDTA-dependent manner than the ferene method on Abbott Alinity *c* (**Figure 1**). In comparison to the other analytes evaluated, much higher concentrations of EDTA are required for a significant decline from baseline, as evidenced by EC50_[EDTA]_ values of 5.6 – 8.86 mmol/L with the ferrozine method (**Table 1**). No significant differences were evident between baseline Fe^2+^ and with up to 8 mmol/L of EDTA when measured by the ferrene method. In the absence of EDTA, the ferene and ferrozine methods generally provided equivalent results (Deming regression, y = 0.98x + 0.10, R = 0.968) in method comparison studies (**Supplementary Figure S5**).

## Discussion

This study arose from an incidental observation of discordant Mg^2+^ results across two analytical methods in a case of EDTA contamination. The scarcity of recent data on the ferene based method performance with EDTA interference, conflicting reports on EDTA effects on Mg^2^□ measurements(1,3,11), and the need for method-specific interference studies further motivated this investigation.

Our findings highlight that EDTA has different effects on Mg^2+^ results depending on the method used. Classically, hypomagnesemia was considered a surrogate marker of EDTA contamination, which was evidentially supported by dye binding methods (1,4–6). However, hypomagnesemia is not a surrogate marker of EDTA contamination when Mg^2+^ is measured with an enzymatic method (**Figure 1**). When using an enzymatic assay, any subtle declines in Mg^2+^ (**Supplementary Figure S1**) due to EDTA are likely to be clinically negligible, which is also supported by two recent studies (2,3) that utilized Abbott Architect instruments. Herein, we have compared both dye and enzymatic methods in the presence and absence of EDTA and have illustrated the striking differences (**Figure 1** and **Table 1**). In contrast to the enzymatic method, when Mg^2+^ is measured by the xylidil blue method, very little EDTA is required to dramatically reduce Mg^2+^ from baseline, with APL_[EDTA]_ ranging from 0.16 – 0.34 mmol/L. The EC50_[EDTA]_ ranged from 0.78 to 1.18 mmol/L when Mg^2+^ was measured by xylidil blue, nearly 1:1 with baseline Mg^2+^ concentrations (**Table 1**). A previous study implicated <0.18 mmol/L of EDTA to cause a 20% decline and 0.92 – 1.83 mmol/L of EDTA to cause >50% decline in Mg^2+^ as measured Roche Modular P-analyzer(12) that also uses the dye binding method. Although baseline concentrations of Mg^2+^ were not reported in the prior study(12), these values align closely with our findings that target a specific decline in concentration by 50% or breach of a specific APL.

It should be noted that dye binding methods are by far the most common method for measuring Mg^2+^. Review of an external quality assurance from the College of American Pathologists indicated that only ∼10% of the participating laboratories were using an enzymatic method in 2021, whereas the top four dye binding methods were those based xylidyl blue (most popular), methylthymol blue, formazan and calmagite (11). While we did not investigate other dye binding methods, these dye binding methods are expected to be affected by EDTA (11,13), although this has not been fully characterized to our knowledge, but postulated based on enzymatic selectivity being greater than both the dye and EDTA for Mg^2+^.

Our study also highlights the method dependent effects of EDTA on Fe^2+^ measurement. From our comparison for Fe^2+^ measurements, the ferrozine method is likely to be less resilient to EDTA contamination when compared to the ferene method. The latter, on the Abbott Alinity *c* shows no significant decline from baseline up to 8 mmol/L spiked EDTA concentration. However, Kalaria et al observed a decline in the presence of EDTA with an Abbott Architect method, notably above 1.86 mmol/L of EDTA and with more than 50% decrease in Fe^2+^ at an EDTA concentration of 5.60 mmol/L(2). Kalaria et al. also concluded that Fe^2+^ should not be reported, but yet indicated that Fe^2+^ did not exceed the wide reference change value and reporting was possibly “debateable”(2). While a similar decline in Fe^2+^ was not evident with our ferene method (Iron2 reagents on Abbott Alinity *c*), the ferrozine method showed notable declines after ∼2 mmol/L EDTA (**Figure 1**). The EC50_[EDTA]_ for the ferrozine method also suggests much higher concentrations of EDTA is required to cause any significant declines in comparison to the other analytes evaluated (**Table 1**).

It was previously suggested that both ferene and ferrozine are affected by EDTA and inhibit colour development(14). While both ferene and ferrozine chromogens bind iron in 3:1 ratio (10,15), the molar absorptivity of the ferene:Fe^2+^ complex is 1.3 times higher than with ferrozine(9). Besides the structural and spectrophotometric differences between ferene and ferrozine, the other differences between the methods include the concentration of ferene and ferrozine, thiourea levels(16), and other possible uncharacterized components that could not be identified. In this study, EDTA in the reaction cuvette was similar in both methods (maximally 0.53 mmol/L and 0.55 with the Roche cobas c303 and Abbott Alinity *c*, respectively), whereas the concentration of ferene was ≥ 1.38 mmol/L versus 0.93 mmol/L of ferrozine.

Both methods for ALP activity and Ca^2+^ measurement illustrated notable declines with EDTA addition (**Figure 1**). When Ca^2+^ was measured by either method, similar EDTA_[APL]_ values were obtained, although EC50_[EDTA]_ were generally greater on the Abbott Alinity *c* method in comparison to Roche cobas c303 (**Table 1**). **Figure 1** suggests that a faster rate of decline in Ca^2+^ is evident from baseline on the Roche cobas c303, although baseline concentrations and patient samples were generally higher by 0.1 mmol/L (or 4.6%) relative to the Abbott Alinity *c* method.

With ALP measurements, there is a notable difference between the two methods (**Supplementary Figure S4**). Despite both claiming traceability to the IFCC reference procedure, baseline concentrations of ALP were higher on the Abbott Alinity *c* but also required higher EDTA concentrations to elicit similar declines (**Figure 1** and **Table 1**).

Based on our assessments of Mg^2+^ and Fe^2+^ measurements on the Abbott Alinity *c*, hypomagnesemia and hypoferremia cannot be used as surrogate markers of EDTA contamination in conjunction with hypocalcemia and low ALP activity.

Standard K_2_EDTA tubes contain 1.8 g/L K_2_EDTA or 6.16 mmol/L, assuming a complete draw. In the scenario where an EDTA plasma aliquot is inadvertently submitted for ALP, Mg^2+^, Ca^2+^, or Fe^2+^ testing, the APL is exceeded for all analytes except Mg^2+^ and Fe^2+^ on the Abbott Alinity *c* (**Table 1**). When between-specimen contamination is suspected, a final K_2_EDTA concentration of 0.2 mmol/L would cause APLs to be exceeded for Mg (Roche) and Ca^2+^ (Roche and Abbott). Depending on blood draw and tube volume, this information provides some insight into differentiation between wrong tube type versus possible tube-to-tube contamination.

This study has several limitations. First, we opted to use Na_2_EDTA instead of K_2_EDTA. However, it was previously shown that the only difference between the two is that Na_2_EDTA does not affect the measured K^+^ levels (12). Second, plasma and serum pools were used to allow for adequate specimen volume, but given the nature of the analytes, no matrix effects or confounding factors are anticipated as a result of pooling material. The specimen pool 3 surpassed the icteric threshold for ALP on Abbott Alinity *c*. However, this pool was retained in our analysis as it was deemed important to include a high concentration ALP sample for the study. An elevated bilirubin in this specimen pool was not unexpected given the ALP elevation. The interference from bilirubin, if any, is deemed minimal based on agreement between the measured ALP concentrations and the Deming regression (**Supplementary Figure S4**).

In summary, while hypocalcemia and low ALP levels are supportive markers of EDTA contaminations, laboratorians need to be aware of the method for measuring Mg^2+^ and Fe^2+^, as hypomagnesmia and hypoferremia are not always markers reflective of potential EDTA contamination.

## Data Availability

All data produced in the present study are available upon reasonable request to the authors

## References

1. Davidson D. Effects of contamination of blood specimens with liquid potassium-EDTA anticoagulant. Ann Clin Biochem. 2002 May 1;39:273–80.

2. Kalaria T, Ford C, Gama R. Managing ethylenediaminetetraacetic acid (EDTA) interference in EDTA contaminated samples - selectivity in reporting analytes. Ann Clin Biochem. 2023 Mar;60(2):92–9.

3. Chadwick K, Whitehead SJ, Ford C, Gama R. KEDTA sample contamination: A reappraisal. J Appl Lab Med. 2019 May;3(6):925–35.

4. Cornes MP, Ford C, Gama R. Spurious hyperkalaemia due to EDTA contamination: common and not always easy to identify. Ann Clin Biochem. 2008 Nov;45(Pt 6):601–3.

5. Lima-Oliveira G, Salvagno GL, Danese E, Brocco G, Guidi GC, Lippi G. Contamination of lithium heparin blood by K2-ethylenediaminetetraacetic acid (EDTA): an experimental evaluation. Biochem Med (Zagreb). 2014 Oct 15;24(3):359–67.

6. Sharratt CL, Gilbert CJ, Cornes MC, Ford C, Gama R. EDTA sample contamination is common and often undetected, putting patients at unnecessary risk of harm. Int J Clin Pract. 2009 Aug;63(8):1259–62.

7. Walmsley TA, George PM, Fowler RT. Colorimetric measurement of iron in plasma samples anticoagulated with EDTA. J Clin Pathol. 1992 Feb;45(2):151–4.

8. Koopman BJ, Hindriks FR, Lokerse YG, Wolthers BG, Orverdijk JF. Injurious effect of EDTA contamination on colorimetry of serum iron. Clin Chem. 1985 Dec;31(12):2030–2.

9. Eskelinen S, Haikonen M, Räisänen S. Ferene-S as the chromogen for serum iron determinations. Scand J Clin Lab Invest. 1983;43(5):453–5.

10. Smith GL, Reutovich AA, Srivastava AK, Reichard RE, Welsh CH, Melman A, et al. Complexation of ferrous ions by ferrozine, 2,2’-bipyridine and 1,10-phenanthroline: Implication for the quantification of iron in biological systems. J Inorg Biochem. 2021 Jul;220(111460):111460.

11. Dent A, Selvaratnam R. Measuring magnesium–Physiological, clinical and analytical perspectives. 2022 Jul 1;105:1–15.

12. Cadamuro J, Felder TK, Oberkofler H, Mrazek C, Wiedemann H, Haschke-Becher E. Relevance of EDTA carryover during blood collection. Clin Chem Lab Med. 2015 Jul;53(8):1271–8.

13. Cacho J, Lopez-Molinero. Angel, Enrique Castells J. Comparative Study of Metallochromic Indicators for Magnesium. Available from: 10.1039/an9871201723

14. Derman DP, Green A, Bothwell TH, Graham B, McNamara L, MacPhail AP, et al. A systematic evaluation of bathophenanthroline, ferrozine and ferene in an ICSH-based method for the measurement of serum iron. Ann Clin Biochem. 1989 Mar;26 (Pt 2):144–7.

15. Hennessy D, Reid GR, Smith F, Thompson S. Ferene - a new spectrophotometric reagent for iron. Canadian Journal of Chemistry. 1984 Apr 1;62:721–4.

16. Narasimhan M, Jaleta K, Adhikari S, Berihun M, SataraNatarajan K, Mahimainathan L, et al. Recent recall of iron reagent-investigation of potential reagent contamination and assay improvement strategy. J Appl Lab Med. 2024 Nov 4;9(6):1040–52.

