## Supplementary Figure S1 for "Assay-dependent Effects of EDTA Contamination on Plasma Magnesium and Iron"

**SUPPLEMENTARY FIGURES**

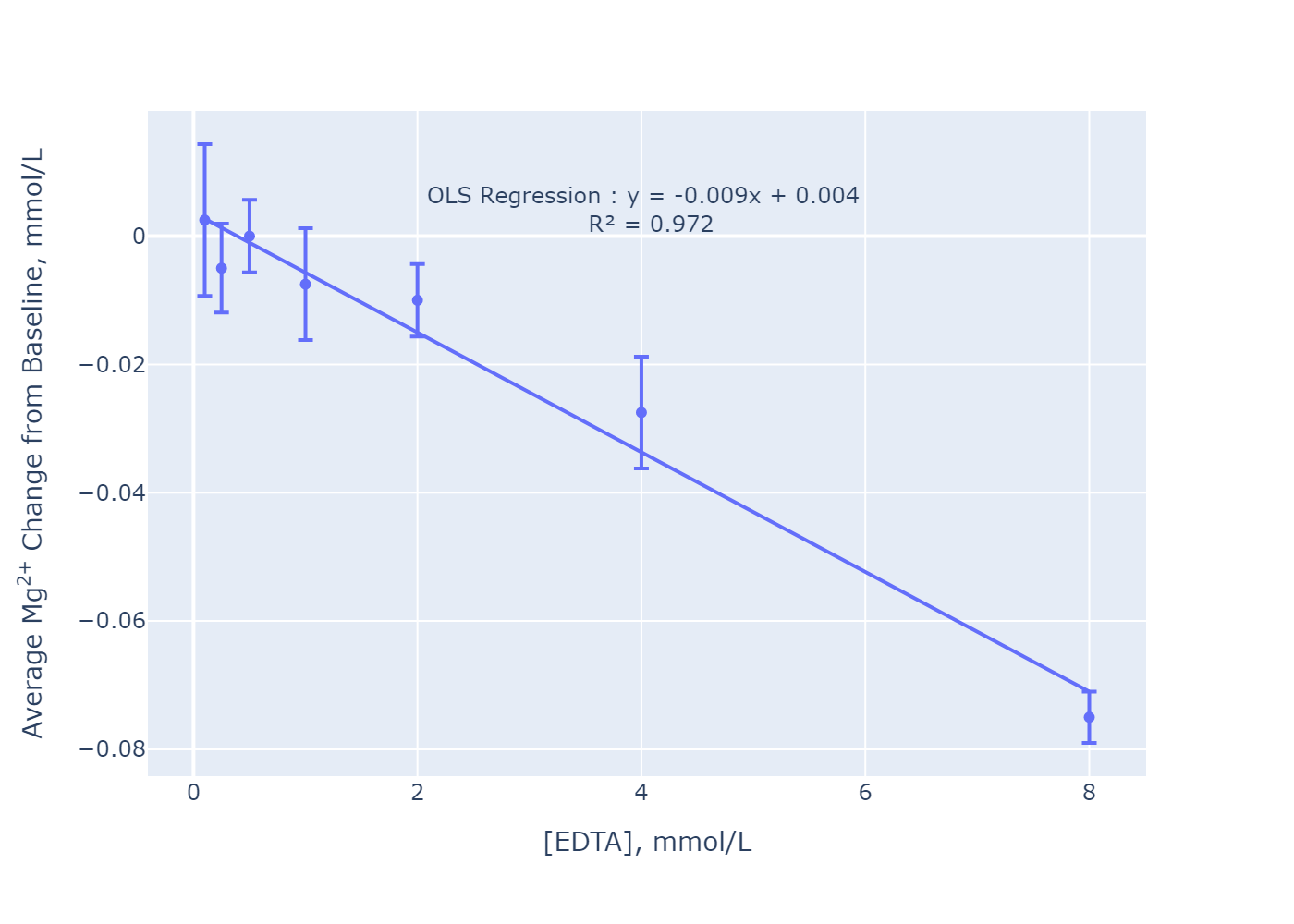

**Supplementary Figure S1:** Average change of Mg2+ as measured on the Abbott Alinity *c*, relative to baseline measurement as obtained across four sample pools as function of EDTA concentration. The average is assessed across the four different sample pools (SP1, SP2, SP3 & SP4) described in the main text. The error bars are based on 95% confidence interval. Linear fitting is based on ordinary least square (OLS) regression.

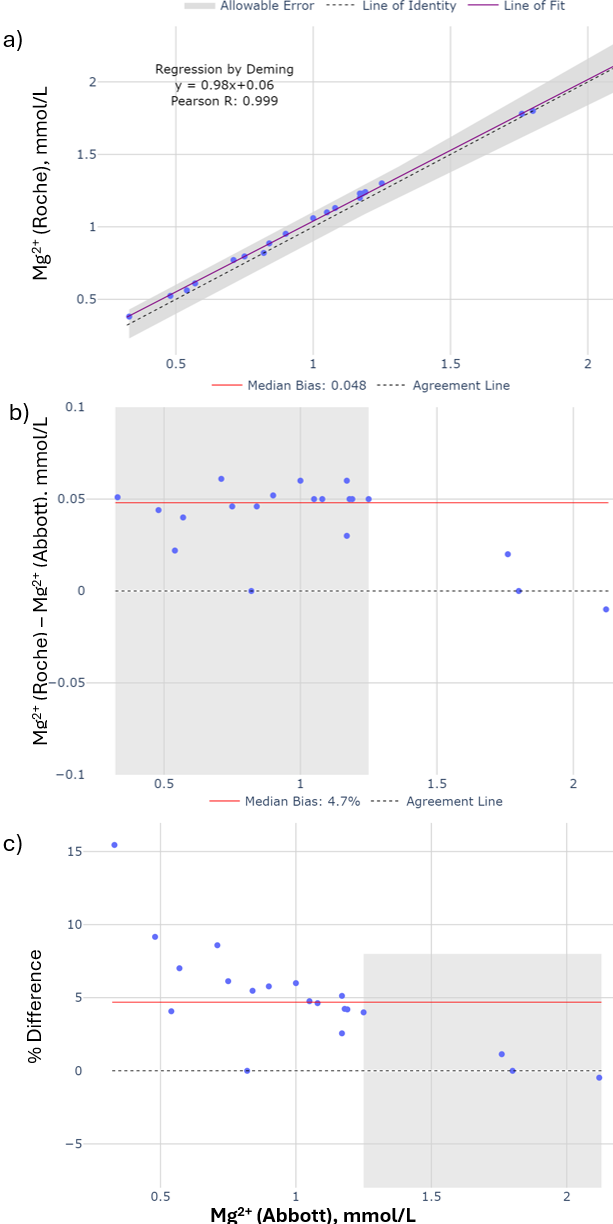

**Supplementary Figure S2:** Method comparison of total Mg2+ on the Abbott Alinity *c* versus Roche cobas c303, with **a)** correlation by regression **b)** the difference plot and **c)** the percent difference plot. Allowable error is indicated by the grey region based on allowable performance limit set by Institute of Quality Management in Healthcare (Accreditation Canada Diagnostics), ± 0.10 mmol/L (at <1.25 mmol/L) and ± 8% (at ≥ 1.25 mmol/L).

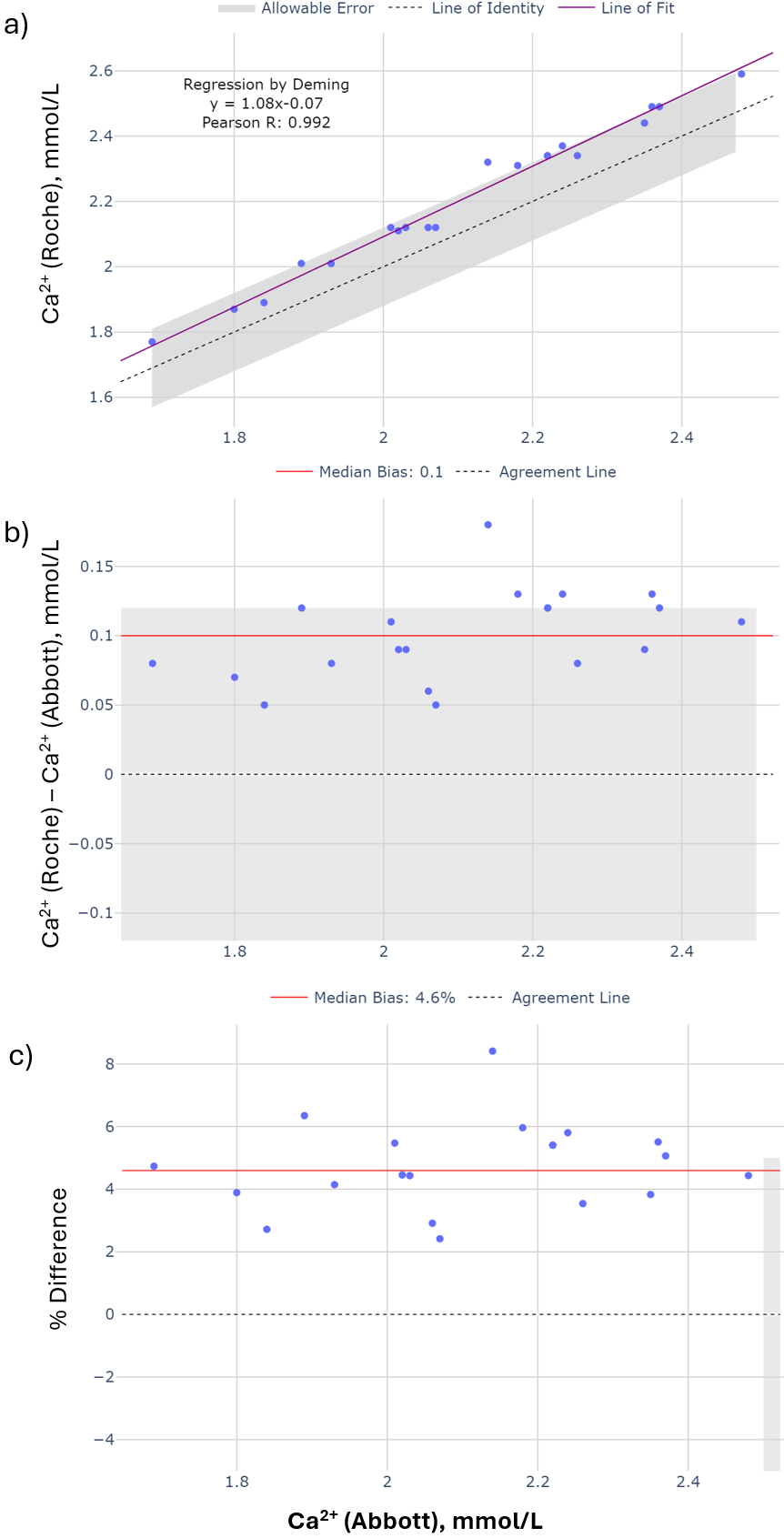

**Supplementary Figure S3:** Method comparison of total Ca2+ on the Abbott Alinity c versus Roche cobas c303, with **a)** correlation by regression **b)** the difference plot and **c)** the percent difference plot. Allowable error is indicated by the grey region based on allowable performance limit set by Institute of Quality Management in Healthcare (Accreditation Canada Diagnostics) ± 0.12 mmol/L (at <2.50 mmol/L) and ± 5% (at ≥ 2.50 mmol/L).

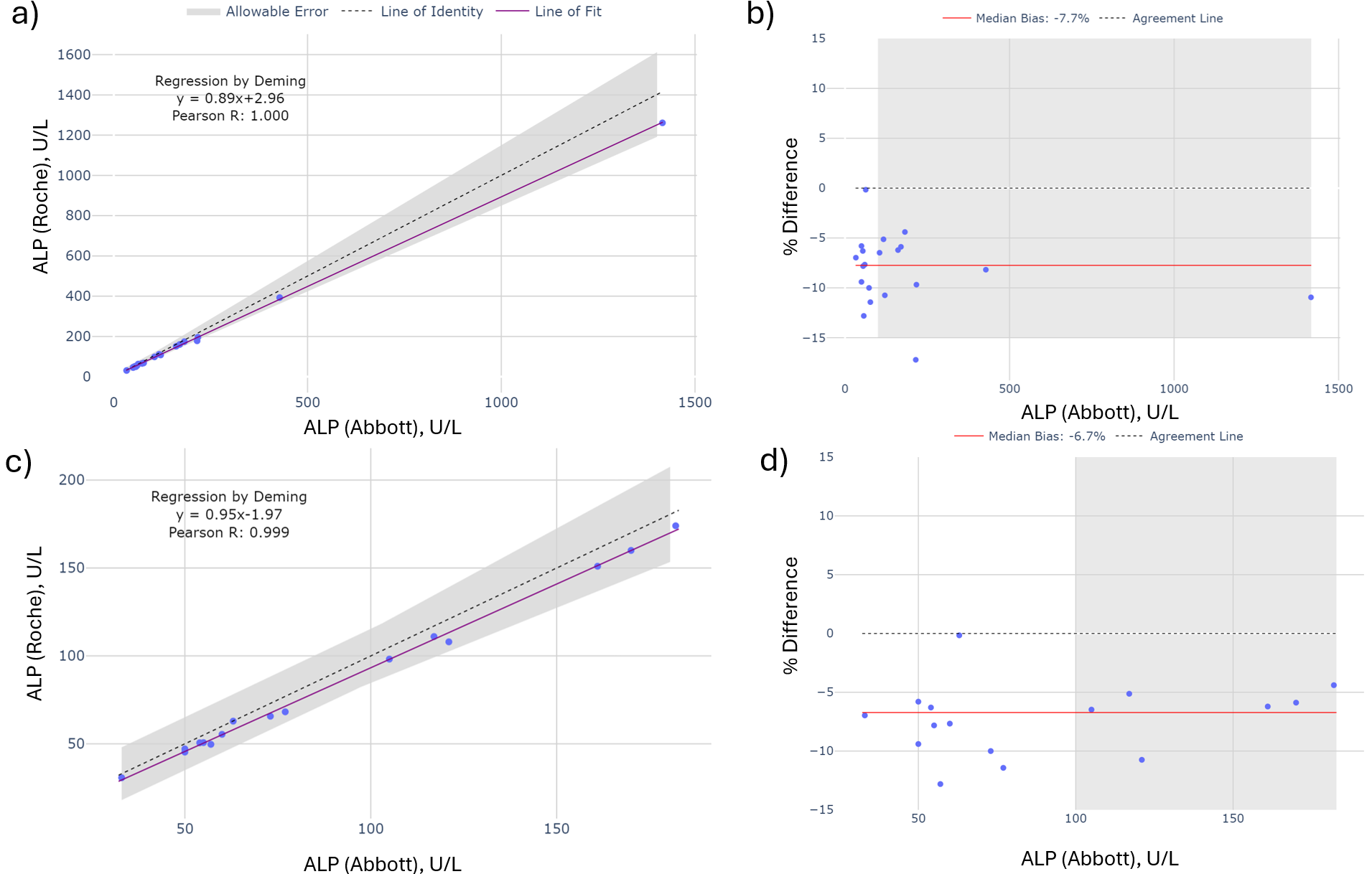

**Supplementary Figure S4:** Method comparison of ALP on the Abbott Alinity c versus Roche cobas c303. Using a full set of 20 samples, **a)** proportional bias is present. The %Difference plot, **b)** suggests a median bias of -7.7% for the 20 samples. The regression plot, **c)** when the highest ALP sample is removed, improves agreement, which leads to a slightly reduced bias, as evident from **d)** the difference plot. Allowable error is indicated by the grey region based on allowable performance limit set by Institute of Quality Management in Healthcare (Accreditation Canada Diagnostics), ±15 U/L when ALP < 100 U/L and 15% when ALP ≥ 100 U/L.

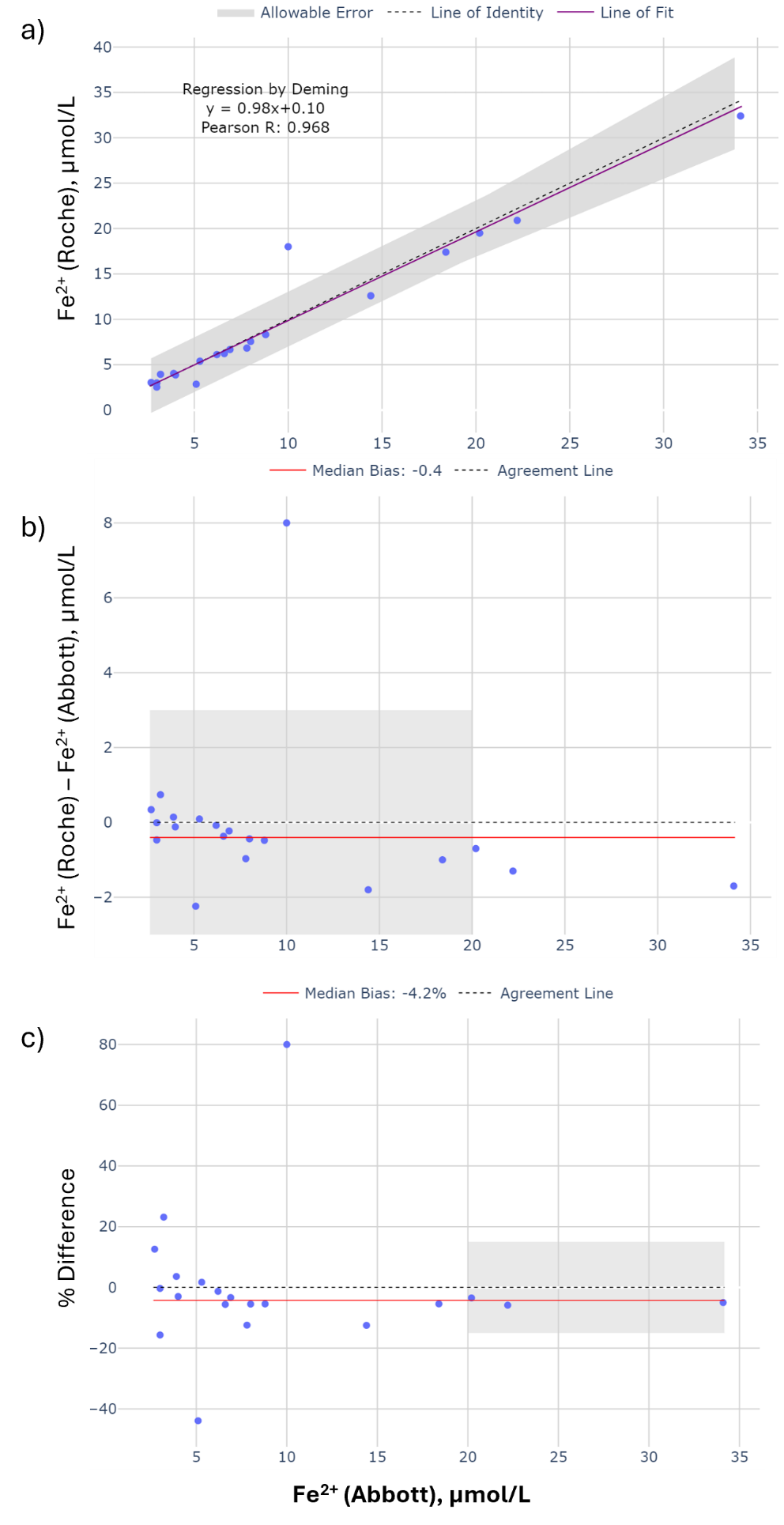

**Supplementary Figure S5** Method comparison of total Fe^2+^ on the Abbott Alinity c versus Roche cobas c303, with **a)** correlation by regression **b)** the difference plot and **c)** the percent difference plot. Allowable error is indicated by the grey region based on allowable performance limit set by Institute of Quality Management in Healthcare (Accreditation Canada Diagnostics), ± 3 μmol/L (at <25 μmol/L) and ± 15% (at ≥ 25 μmol/L).
